# Stones in Our Pockets: Mental Health Dimensions of Grief in Contemporary Video Games

**DOI:** 10.1101/2024.04.23.24306235

**Authors:** Emma Reay, Minhua Ma, Anna Mankee-Williams, Gabriela Pavarini, Nicola Shaughnessy, Kamaldeep Bhui

## Abstract

This article combines the expertise of games studies scholars, medical ethicists, and clinical psychologists to analyse representations of grief in contemporary video games. Grief is a universal human experience, but navigating its psychological, social, and embodied effects can be a challenging task. Constructionist approaches to grief therapy emphasise the importance of metaphors for understanding the experience of loss (Nadeau 2006; Young 2008; Neimeyer 2010; Southall 2013). Paying attention to the metaphorical language used by a client can provide a therapist with key information about the client’s personal and cultural perspectives and world view. Equally, a therapist can work with clients to devise metaphors that shift their perspectives and aid the process of meaning-making. Video games provide players with new metaphors to express and explore grief. Since video games communicate across audio, visual, verbal, haptic, mechanical, and performative modes, they can offer a complete gestalt that touches on the physical, emotional, practical, and systemic impact of loss. In this article, we survey the multimodal metaphors for grief that appear in 14 commercial-off-the-shelf (COTS) video games and identify recurring tropes and themes. We consider 1) what is illuminated by these metaphors? 2) what is obscured by these metaphors? and 3) what are the therapeutic implications of these metaphors? We conclude with a set of recommendations for game developers who want to design ‘serious games’ that explore emotionally fraught topics, and a set of recommendations for grief and bereavement therapists seeking to integrate video games into their practice.

## Introduction

Grief is an unavoidable part of human experience, but navigating its psychological, social, and embodied effects can be a difficult task. While most people find ways of living with grief, approximately 10-15% struggle with “intense, prolonged and complicated grief, characterized by extreme separation distress, preoccupation with the loss, and the inability to function in major life roles across a period of many months or years” (Neimeyer & Thompson, 2014). Over the last two decades, there has been a rise in the recognition of the importance of metaphors in psychotherapy (Lawley & Tompkins 2011; Southall 2013), including a growing awareness of how metaphors exchanged between therapists and bereaved clients might affect attitudes towards grief (Nadeau 2006; Young 2008; Neimeyer et al. 2010; Goldberg & Stephenson 2016; Guité-Verret et al. 2021). Psychotherapists have drawn on the work of linguists and literary theorists such as George Lakoff and Mark Johnson (1980), Paul Ricoeur (1975), and Susan Sontag (1978), to argue that metaphors buttress our entire conceptual systems, affecting how we think, talk, and act (Southall 2013). Metaphors are not mere descriptions of reality: they are the means by which reality is negotiated, both on an intra-psychic and a societal level.

Metaphors can be useful in therapy because they ground abstract, intangible, ephemeral experiences, making them easier to examine and easier convey to others. In other words, metaphors are an aid both to reflection and to communication. At the same time, metaphors are inherently polyvalent. They are laden with intended and unintended connotations, which means they are somewhat resistant to the reduction in complexity that occurs when lived experience is translated into language. Furthermore, metaphors can provide a “safe way” (Nadeau, 2006 p.220) to explore difficult emotions or uncertain futures because they allow patients and therapists to approach sensitive topics indirectly. This can reduce defensiveness and instead encourage curiosity, flexibility, and even playfulness.

Finally, metaphors can reveal what is being obscured or ignored. Since metaphors explicitly offer a subjective perspective on an experience, they can throw into sharp relief possible alternative perspectives that can subsequently be examined.

To support the generation and integration of metaphors, therapists and patients often engage with literature, art, and performance. This can take the form of creating new literary, artistic, or performed pieces, or of making personal connections to existing pieces. The process of exploration, articulation, validation, and transformation that structures arts-based therapy can be particularly useful for those experiencing bereavement. This is because grieving can be understood as a process of reconstructing a world of meaning that has been undermined by loss. To put it another way, grieving is an act of sense-making that allows one to repair or redefine one’s model of reality. Traumatic experience can disrupt previously held perspectives through impacts on memory but also by fracturing moral frameworks by which people live their lives.

Making art or engaging with art can provide the scaffolding within which one can build new frames of reference for interpreting one’s experiences. Arts-based therapy can engage with a wide range of media, such as photography, dance, pottery, and poetry. Video games are a relatively new medium, but nonetheless hold an important position in contemporary media landscapes. They appeal to overlapping, adjacent, and different demographics compared to traditional media, thereby expanding the reach of arts-based therapies. It is possible that patients who are unwilling to engage with traditional media such as poetry or painting could be more open to gaming (Pavarini, Reay, & Smith).

A poignant example of using metaphor in therapy is found in narrative exposure therapy, where stones can represent traumatic events or profound sadness, such as grief, in a person’s life timeline. Each stone, varying in size and weight, symbolises the magnitude and impact of different traumatic experiences. This physical representation allows individuals to organise their life stories visually and tangibly, acknowledging and addressing each traumatic event. Through this process, patients can explore and understand their emotional landscapes in a structured, tangible way, facilitating healing and comprehension of their journeys. This method exemplifies how metaphors in therapy can bridge the gap between the ineffable complexities of personal experiences and the necessity of addressing them directly for therapeutic progress.

While there have been important studies that address the use of video games in therapeutic settings (e.g., Ceranoglu 2010), gaming as a form of self-care (e.g., Spokes 2024), table-top role playing games in group therapy (e.g., Connell 2023) and game-making in relation to bereavement (e.g., Harrer 2018), there is a need for further empirical and theoretical research into how this new medium fits within existing therapeutic paradigms (Pavarini, Reay, & Smith). This article contributes to current scholarship in three ways:

1. It integrates insights from games studies about how metaphors work in video games and considers the connection to how metaphors work in psychotherapy.
2. It examines metaphors related to grief across a wide range of video games, from small, browser-based or mobile games developed by solo designers to AAA console games by major development teams.
3. It models a form of interdisciplinary research wherein psychologists and psychiatrists can comment on the therapeutic value of commercial video games, without requiring extensive gaming expertise.

## Literature Review

### Metaphors in games

Metaphors drawn from video games and gaming culture are already shaping personal behaviours, societal attitudes, and cultural beliefs. Describing someone as an ‘NPC’ (non-player character), for example, proliferated as a meme in far-right political circles to ridicule those who did not hold conservative views. Similarly, video game specific concepts such as ‘side quests’, ‘the final boss’, ‘noobs’, ‘XP’, and ‘rage quitting’ are now used as figures of speech in non-gaming contexts. This paper, however, is interested in the use of metaphors in video games and how these metaphors might influence conceptions of grief.

Video games provide us with new languages to express and explore complex aspects of the human experience. While we should be wary of contributing to the rhetoric of ‘video game exceptionalism’ by overstating the novelty of this medium’s affordances (Nicklin 2020), video games do combine communicative modes in ways that differ significantly from other forms of entertainment. Firstly, as Harrer writes, “unlike other artistic methods…game design engages metaphors through multiple modalities at once: rules, haptics, graphics and sounds” (2018, p.62). This means that video games can simultaneously draw on:

- the haptic, somatic languages of dance and performance,
- the visual imagery of film, animation, and photography,
- the auditory devices of music, soundscapes, and sound design,
- the verbal modes of dialogue, narration, and written text,
- the systematic modelling of simulations,
- the ludic signifiers of play, puzzles, challenge, and competition.

Reay (2024) argues that video games can be understood as dynamic ecosystems of meaning, within which different elements like visual, auditory, verbal, haptic, and ludic can enhance, extend, counterpoint, and even contradict each other. Dissonance between these elements can create interpretive gaps that invite subjective interpretation. The multimodality of video games does not necessarily close interpretive gaps, thus rendering a simulation of reality.

Instead, introducing various elements creates spaces for interpretation, enriching the game with complexity and multiple perspectives. While aiming for ultra-realism in depicting loss could potentially re-traumatise players, using these elements to deepen and add subtlety to metaphors empowers players to construct their own meanings. This approach leans into the imaginative rather than just the factual and literal representation, trusting players to close interpretive gaps. Additionally, since video games typically encourage playful engagement and exploration, players might see these open-ended interpretations as challenges to be unraveled.

Secondly, the highly interactive nature of video games means that they offer players a unique set of possible subject positions (Thi Nguyen 2020). In relation to video games that use extended metaphors, Magnusson notes that players are invited to respond (cooperatively or antagonistically) to the game whilst also being ‘ventriloquised’ by the game (2022). He claims that this form of “triangulated address and poetic ‘ventriloquism’ can only really be achieved by interactive media” (p.72, 2022). This is affirmed by Harrer, who notes, “compared to other modalities of reception, [video game] play invites the receiver to become part of a symbolic world, allowing an embodied connection to personal contents and themes” (p.63). Players can be interpellated into metaphorical representations, discovering how they work and what they feel like from the inside out.

Rusch frames this as the “crucial difference between *doing* and *witnessing*” (p.72), coining the term ‘experiential metaphors’ to capture this distinction. Experiential metaphors are “a powerful form of metaphorical mapping and meaning generation” wherein inner states are made “concrete by letting us physically enact them through our virtual bodies or other means of projection into a game environment” (p.74). This is important because it engages ‘the emotions of agency’ (Isbister 2008), which include feelings such as pride, guilt, triumph, betrayal, humiliation, complicity, gratitude, loyalty, and duty. Unlike emotional reactions that we might experience vicariously via empathy with on-screen characters, the emotions of agency are felt ‘first hand’ and belong solely to the player. This contributes to the sense that a particular playthrough is personal and reflects a player’s individual choices, desires, and competencies, which may prove particularly useful in the context of deepening self-awareness and re-writing one’s identity in the aftermath of loss.

Finally, video game metaphors stand out from those in other media by offering visual metaphors and non-verbal communication that can intuitively reach and resonate with the player, often more directly than the more cognitively demanding verbal metaphors. Metaphors are, on some level, fundamentally about the connections between different elements since they invite us to understand one thing in terms of another. Video games take this a step further by specifically emphasising the connections between contexts, inputs, and outcomes. This could be particularly useful for encouraging bereaved patients to locate their personal experiences within a network of meaning, or – to use a metaphor commonly associated with grief – to see the hole created by loss within the context of the remaining material that edges it. As Rusch summarises, taking a systemic view of “personal problems can be highly beneficial” because it can “open one’s perspective to the bigger picture rather than focusing on an issue in isolation” (p.171). This becomes even more important when we consider Frazer-Carroll’s critique of the individualisation of mental illness. She writes that the failure to locate individual suffering within a broader social and political matrix “frames ongoing distress as a personal failure to self-discipline or seek out appropriate services, rather than acknowledging the structural conditions that also dictate our lives” (2023, p.44). Representations of loss via systems-based media may challenge the ‘privatisation’ of loss (Jalland 2013), instead emphasising the communal, structural, and ritual aspects of grief.

### Video Games and Arts-based Therapy

We are not the first to suggest that video games can be used as part of arts-based therapeutic interventions. Beyond the ‘games on prescription’ model – which has seen purpose-built video games such as EndeavorRX and LumiNova approved to augment the treatment of ADHD and anxiety respectively (Waltz 2020; Lockwood et al. 2022) – there has been increasing scholarly interest in the therapeutic potential of playing Commercial-Off-the-Shelf (COTS) games (Phillips 2021), as well as in the therapeutic potential of amateur game-making (Danilovic 2018; Harrer 2018; Rusch 2014).

We have chosen to focus on the representation for grief in COTS games rather than to analyse custom-made serious games (defined here as games made primarily for a purpose that is beyond entertainment) because we are interested in how the artistic or literary quality of video games might shape grief management. While some serious games for grief have a fantasy layer - the mobile game, *Apart of Me,* for example invites players to explore a tropical island whilst completing therapeutic tasks such as breathing exercises and logging one’s mood - they are often better categorised as ‘tools’ rather than as ‘art’. That is to say, the success metrics of a video game like *Apart of Me* are connected to its instrumental value, rather than to its artistic value. The fantasy layer exists to make the core therapeutic tasks more palatable: the audiovisual and narrative components are the ‘chocolate’ in the ‘chocolate-coated-broccoli’ model of serious games. Our view, however, is that the figurative aspects of video games could be considered ‘nutritious’. If we accept that it is the affective, imaginative, and aesthetic aspects of poetry, novels, art, sculpture, and so on that have the power to clarify grief and comfort the grieving (Shapiro 2006; Brennen 2020), then it follows that we should explore these same aspects in video games used in therapeutic settings for grieving patients. Finally, unlike custom-made serious games for therapeutic interventions, COTS games often have large player bases and appeal to broad audiences, which may include grieving individuals. Arguably, the reach and penetration of COTS games may make their impact on collective attitudes towards grief more significant than serious games.

### Surveying Metaphors used in Grief Therapy

Researchers have documented the metaphors used by clients to describe their grief in the form of case studies (Young 2008; Goldberg & Stephenson 2016; Guité-Verret et al. 2021). Young looks for categories that emerge within her case studies and identifies two groups of metaphors – “one connotes breakage, the other erasure” (2008, p.361) – and three themes – “metaphors of natural forces, of technology, and of fiber and fiber arts” (p.363). Guité-Verret et al. identify three distinct categories of metaphor in their case studies of individuals experiencing bereavement during the Covid-19 pandemic: “1) being cut off from others, 2) living and facing obstructions and 3) feeling shockwaves” (2021).

Other researchers have conducted narrative literature reviews to identify the most common metaphors for grief reported in sessions with therapists. Southall (2013), for example, concludes that the dominant metaphors for grief can be sorted into the following categories: military metaphors, metaphors of journeying, metaphors of personhood, metaphors of the natural world, and existential / ideological metaphors.

Rather than reporting metaphors used by patients and therapists to describe grief, this paper reports the metaphors used in contemporary video games to describe grief.

However, we, too, are interested in cataloguing and critiquing the emerging motifs and patterns.

Based on this literature review, we have generated the following set of assumptions:

1. Metaphors are important meaning-making tools for those experiencing grief. Metaphors can provide patients and therapists with insights into subjective, personal experiences by both illuminating and obscuring aspects of reality.
2. Video games employ metaphors in ways that are comparable to other art forms and entertainment media. However, the form and function of metaphors in video games are shaped by a combination of communicative affordances that is unique to this medium.
3. Video game players may encounter metaphors for grief in COTS games that have an impact on their own mourning processes.

## Method

We employed purposive sampling to select video games for this study. Our inclusion criteria were:

1. Commercial off-the-shelf games (rather than bespoke games specifically designed as a therapeutic intervention)
2. Published between 2013 and 2023
3. Accessible on mobile, PC, Nintendo Switch, or PlayStation
4. Available to play in English
5. Represents a form of grief (e.g., death-related loss, relationship dissolution, losing home/possessions, loneliness, or major life transitions)

Table 1 details our final corpus.

**Table 1.** Selected video games analysed for grief representation.

|  | **Title** | **Publication Year** | **Platform** | **Type of Grief** |
| --- | --- | --- | --- | --- |
| 1 | Florence | 2018 | Mobile | Relationship dissolution |
| 2 | Spiritfarer | 2020 | Nintendo Switch, mobile | Bereavement |
| 3 | That Dragon, Cancer | 2016 | PC | Bereavement |
| 4 | Gris | 2018 | PC | Major life transition, bereavement |
| 5 | Stillness of the Wind | 2019 | mobile | Age, death, loneliness |
| 6 | Far From Noise | 2017 | PlayStation, mobile | Life and death, helpless, loneliness |
| 7 | Brothers: A Tale of Two Sons | 2014 | PlayStation | Bereavement |
| 8 | God of War | 2018 | PlayStation | Bereavement |
| 9 | If Found... | 2020 | Mobile | Losing home, major life transition |
| 10 | Röki | 2020 | PC | Bereavement |
| 11 | Knights and Bikes | 2019 | PC | Bereavement |
| 12 | Lost Words: Beyond the Page | 2020 | PC | Bereavement |
| 13 | What Remains of Edith Finch | 2017 | PC | Bereavement |
| 14 | Under a Star Called Sun | 2020 | PC | Bereavement |

Our corpus includes AAA games (e.g., *God of War*), award-winning games (e.g., *What Remains of Edith* Finch), indie games (e.g., *Spiritfarer, Lost Words: Beyond the Page*), autobiographical games (e.g., *That Dragon, Cancer*), and single-developer games (e.g., *Under a Star Called Sun*). It encompasses action games, walking simulators, platformers, adventure games, visual novels, cooperative games, and resource management games. It reflects a range of art styles, from high fidelity, motion-captured, realism through to highly abstract, stylised, pixel art. Finally, our corpus contains games that might appeal to a diverse range of player demographics, including different ages, genders, and levels of gaming literacy.

### Autoethnographic Reading

Autoethnographic close reading is an interpretive approach that involves analysing the formal properties of videogames while considering the role of the player in the signification process. According to Kłosiński (2022), autoethnography requires the transformation of lived experiences into discursive ones through narration, which can be compared with other narrations about it. In the context of the formal analysis of video games, autoethnographic reading requires the player-analyst to document their gameplay experience. The resulting text aims to capture the expressive eloquence of a video game’s non-verbal signifiers, as well as the embodied, affective experience of play (Reay 2024). The point of autoethnographic reading is not to create an objectively accurate account of a specific gameplay sequence, but to preserve the unique aesthetic and emotional journey of an individual playthrough. These written accounts can be used as stable reference points throughout the analytic process.

For this study, the lead author played all of the video games listed in Table 1, creating anecdotes that detail key aspects of each playthrough. These narratives encompass the visual, auditory, mechanics, and haptic dimensions of the grief metaphors found in each game. This initial summary was then refined in collaboration with co-authors, who each experienced at least one game from the selection. Through hybrid meetings and workshops, the team identified points of resonance and dissonance based on their individual play experiences. These discussions helped organise their observations and reflections, leading to the selection of theoretical frameworks for analysing the games. This approach provides a subjective perspective for game analysis. The collective insights were integrated with the team’s expertise in serious games, arts-based therapy, and medical ethics, fostering a comprehensive and interdisciplinary exploration of the games and their potential implications.

It is often assumed that interdisciplinary collaboration enhances research outputs. However, we have found that studies located at the intersection of psychology and games studies often encounter barriers that inhibit productive knowledge-exchange and knowledge-creation. Some of these barriers are general and relate to a mismatch between humanities methods of enquiry and methods used in the social sciences, but others are specific to the study of video games and relate to levels of ludic literacy. Ludic literacy refers to 1) a proprioceptive fluency when interacting with conventional control schema for video games; 2) the ability to ‘read’ a video game’s audiovisual layer – including icons and symbols - to interpret its underlying rules; 3) a familiarity with the formal properties of different game genres. Asking a researcher with low ludic literacy to appraise a video game is akin to asking someone who cannot read to critique a novel. This is the elephant in the interdisciplinary room: ludic literacy is not considered a skill in the same way that traditional literacy is considered a skill, and so ludic illiteracy often goes unremarked.

We feel it is not necessary for those with expertise in psychology and psychiatry to also be experienced gamers to contribute productively to discussions about video games in therapeutic settings. It is the role of games studies scholars to make the primary texts legible to collaborators. This is usually an iterative process that requires designated space and time. For this project, we opted to establish this collaborative space through virtual or hybrid meetings.

## Results

**Table 2.** Summary of grief narratives, visual, auditory, game mechanics, and haptic features in selected games.

| Title | Grief Narrative | Visual | Auditory | Game Mechanics | Haptic |
| --- | --- | --- | --- | --- | --- |
| **Florence** | A romantic relationship comes to an end. | The vibrancy and variety of the colour palette represents levels of happiness and fulfilment. Blues and greys are used to represent the lowest point in the protagonist's experiences, warm yellows represent the love interest, and pinks and greens signal the protagonist's recovery. | This wordless game relies on a piano and a cello to 'give voice' to the romantic partners. When the protagonist loses her lover, the cello shifts into a minor key and plays a sad dirge, while the piano becomes quiet, fragmented and unsure. | Attempting to reassemble a torn photograph of the lovers while the pieces drift slowly but inexorably off the edge of the screen. Attempting to complete a jigsaw puzzle of the lovers only to discover that their adjoining pieces are incompatible. | Tapping repeatedly conveys emotional effort – for example, getting the protagonist to get out of bed in the nadir of her depression requires 20 taps of the same button. Swiping and dragging conveys efforts to maintain 'wholeness' in the face of fragmentation. Refraining from touching the screen expresses the counterintuitive passivity and patience required to 'move on' after a breakup. |
| **Spiritfarer** | A young, female captain ferries the souls of the dying towards death. Eventually, the captain must ferry herself towards death. | Souls initially appear as shrouded, anonymous green ghosts. Once aboard the boat, they take the form of anthropomorphic animals. Death is represented by 'the Everdoor', which is a cherry blossom strewn bridge above a red river. The animals disappear in a burst of golden light and reappear as a constellation of stars in the night's sky. After death, the animals' homes remain empty on the boat and a symbolic flower is found on their bed. | The game is accompanied by a soothing but melancholy soundtrack that features piano and acoustic guitar. The same soaring, gentle theme music plays as each spirit passes through the Everdoor. | Palliative care takes the form of farming, cooking, weaving, listening to personal stories, and dispensing hugs. Mining logging, and trading goods are also key mechanics as part of the game's resource management system. Players explore the ocean to pick up souls, uncovering new areas of the map in the process. | While this game does contain some light platforming elements and some timed challenges, the controls are simple and streamlined. There are plenty of lulls in productivity wherein the input controls are simply clicking through boxes of dialogue. The steady day-to-night cycle of gameplay encourages a familiar rhythm of movements, giving the input gestures a ritualistic feel. |
| **That Dragon, Cancer** | A family navigates their young son's terminal | The low poly visuals are often surreal, and the game cuts suddenly between images and scenes. | As this is an autobiographical game, the voices of the children are taken from | Steering Joel through outer space, dodging the spiky cancer cells that threaten to pop his chosen form of | The input controls are somewhat limited, but the camera control via the mouse is very open and free. This places |
|  | diagnosis and death. | A beautiful island with green spaces and a playground suffers from an incursion black brambles and spiky dead trees. Different versions of a faceless child (baby Joel) play on the swings, slide, and climbing frame. The black brambles pulse and multiply, and then Joel is seen laying on a stretcher wired up to a heart monitor. These black brambles later appear in the hospital, wrecking Joel's treatment room.<br>As Joel's parents receive the news of his terminal diagnosis, the room slowly fills with water. As Joel's condition deteriorates, his father 'drowns' in an ocean filled with messages-in-bottles, while the mother drifts calmly in a rowing boat with no oars. | the family's home videos. This is especially moving when it is the sound of Joel's laughter or of his crying. | transport – a bunch of balloons made from inflated surgical gloves. Joel celebrates final treatment day with a rally cart race around the hospital corridors. The laps are measured in weeks and years and the collectible power ups are chemotherapy and pain-management drugs.<br>A game-within-a-game, an arcade platformer where Joel flees an undefeatable dragon. | a lot of importance on the idea of seeing through someone else's eyes and experiencing multiple viewpoints. |
| **Gris** | A girl traverses a barren, faded landscape, gradually triggering colour to return. | The world begins grey. As the avatar journeys through the world – and through the five stages of grief - hand painted water colours are layered over the environment: numb grey for denial, arid red for anger, hopeful green for bargaining, rainy blue for depression, hints of gold for acceptance. Inky black leaks into every environment, occasionally overpowering all other colours. Importantly, these colours do not replace each other, but blend to form a richer and beautiful image. | Orchestral score with angelic vocalisations. Initially, the protagonist loses her singing voice and ends up in a nearly silent world. At the game's conclusion, she finds her voice again and sings the statue of the woman back together. She sings until her last breath, but is overcome by the waves of lightless black, | A new ability is introduced for each stage of grief:<br>Anger – you can become a sturdy cube that allows you to smash things and also withstand the harsh, red wind that blows you backwards.<br>Bargaining – you collect apples for a friendly, little robot who ultimately leaves you. | In the first level (denial), the player must apply constant pressure to the analogue stick to keep the protagonist slowly moving forward. Otherwise, she collapses to her knees.<br>Since opportunities to interact with the delicate environment are limited, smashing things using the 'cube' ability feels rewarding and cathartic.<br>Fleeing persecuting shadows creates feelings of panic, and navigating the intricate caverns |
|  |  | Statues of women fill the gameworld, either functioning as support pillars in temples, with their hands covering their weeping faces, or crumbling to dust in ruins.<br>The collectibles are small, glowing white orbs that resemble fireflies but leave geometric trails behind them that evoke astrological constellations. A repeating visual motif is trees as neurons as circuit boards.<br>Another motif is asymmetric shadows and reflections, which sometimes become antagonistic. For example, the shadow of a murmuration of many little birds that usually help the avatar to jump suddenly accumulates into one huge bird that chases the avatar. | which seeps into her mouth. After a beat, the statue sings back to her and dispels the blackness. The statue and the girl sing a harmonising duet. | Depression – swimming through the darkness in search of lights.<br>Acceptance -You can flip perspective and turn ceilings into traversable floors.<br>There are ‘achievements’ to unlock. In the first level, for example, if you refuse to follow the conventions of a side-scroller and instead go back to the ‘beginning’ (the far left-hand side), the protagonist meets with a crumbled statue and is awarded ‘denial’. | of ‘depression’ is confusing and often leads the player to disappointing dead ends. |
| **Stillness of the Wind** | Explores themes of age, death, and loneliness through the daily life of an elderly woman, Talma, tending to her farm in a desolate desert. Players encounter the quiet grief of isolation and the looming presence of death through<br>her daily routines. | Serene, minimalist visuals that capture the stark beauty of Talma's desert farm. The game's art style, with its muted colour palette and simple, elegant design, evokes the solitude and tranquility of rural life, enhancing the game's themes of isolation and reflection. | a subtle auditory landscape, emphasising ambient sounds like the wind's whisper, animal noises, and the quiet rustle of nature. This minimalist sound design complements the game's themes, reinforcing the sense of solitude and the poignant atmosphere of Talma's isolated existence. | The game employs mechanics similar to casual farming games, with a distinctive twist: its pace gradually slows, symbolising Talma's aging. This mechanic not only adds depth to gameplay but also reflects the themes of time's passage and the inevitability of change, enhancing the narrative's exploration of life, death, and loneliness. | The haptic aspect is intuitive and straightforward, mirroring the simplicity of the game's farming mechanics. Players interact with the game objects and manage tasks through uncomplicated inputs, with the game's pace intentionally slowing to reflect the protagonist's aging. |
| **Far From Noise** | The player is trapped in a car on a cliff's edge, conversing with a deer about life, death, helplessness, and loneliness, encouraging reflection on existence and grief | The game features stunning 2D visuals, with the game's set piece—a car perched precariously on a cliff's edge—serving as a backdrop for the unfolding narrative. The visuals transition from day to night, capturing the serene beauty of the natural environment and enhancing the game's contemplative mood. | Marked by its natural ambiance, with the sounds of nature and wind predominating. The absence of a traditional music score emphasises the game's serene and contemplative atmosphere, immersing players in its tranquil setting. | Game mechanics centred around dialogue options, allowing players to shape the protagonist's life story. Through these interactive elements, players can decide on aspects like the subject she studies and the reason for leaving college, thereby influencing the narrative's direction and depth. | minimalistic haptic, primarily on selecting dialogue options through simple inputs. This approach aligns with the game's narrative-driven nature, ensuring that the player's engagement is more about making choices and less about complex physical interactions. |
| **Brothers: A Tale of Two Sons** | Two brothers set out on a quest to retrieve an elixir that will save their dying father. On route, the older brother is fatally wounded. | Grief is expressed through the characters' body language. The dead mother also appears to Little Brother as a translucent spectre at certain points in the game. | After his death, Big Brother's voice is heard during a moment of desperation whispering soft words of encouragement to his brother. | Loss is evoked through the sudden shift from a co-operative set of mechanics (with the two brothers using their unique skills and abilities to solve environmental puzzles together), to limited single-player mechanics that make the player feel vulnerable, disoriented, and underpowered. | The player directs Big Brother with the left hand and Little Brother with the right hand. After Big Brother's death, the left hand becomes redundant, and all the buttons previously associated with this character no longer work. Later on, in a moment of desperation, the left-hand controls briefly reanimate, giving Little Brother the strength he needs to complete the game. Immediately after Big Brother's death, Little Brother has to bury him. The analogue stick requires constant pressure to keep Little Brother slowly moving to complete the task or else he collapses to the ground immobile. |
| **God of War** | A god with a violent past is accompanied by his pre-teen son on a pilgrimage to scatter his recently deceased wife's ashes. | The environment is marked with golden paint, which indicates the path forwards (e.g., which ledges are climbable etc). The gold colour is associated with the dead mother. The idea that she has left this trail for her husband and son is validated at the end of the narrative, where her prophesying abilities are discovered. As Kratos finally embraces his new role as 'father', he allows the bandages covering the chain-shaped scars on his forearms to be blown off and permits his son to carry the ashes. | Fully voice acted characters. Kratos is gruff and tight-lipped, but his son often expresses his grief aloud, with emotions ranging between sadness, anger, and resignation. A woman is heard singing in an ancient Scandinavian language at the game's conclusion. | Kratos fights using two weapons: a Frost Axe that belonged to his dead wife, and the fiery Blades of Chaos, which represent his previous life as a bloodthirsty God of War. Read metaphorically, the ice is aligned with Kratos' repression and denial, while the blades are aligned with his rage. In contrast to Kratos' constant, brutal death-dealing, his son can be directed to read glyphs inscribed on ancient slates that relate aspects of Norse mythology. His aptitude for languages and communication is linked to his mother, who first taught him the Norse stories. | The combat is very 'juicy', which gives gameplay a cathartic feel. In contrast, there are lulls in action when Kratos rows himself, his son, and his sidekick Mimir around a flooded realm. During these calm moments, dialogue is exchanged that provides exposition, deepens character relationships, and furthers the plot. |
| **If Found...** | A young transwoman, Kasio, experiences rejection and isolation upon her return to rural Ireland. | The abandoned house that the group of young queer people squat in is slowly decaying and becoming uninhabitable. A sci-fi subplot revolves around a black hole that is going to consume the world. A female astronaut called Cassiopeia liaises with a ground control agent to try to avert the disaster. | In the moments before Kasio's near death, the electronic synth soundtrack falls silent. As she succumbs to a fever, the game switches to the subplot of the blackhole and shows worlds and galaxies collapsing into | The single core mechanic in this game involves erasing pages of a journal to reveal the subsequent plot beat. After Kasio's near death experience and her reconciliation with her family, the eraser becomes a pencil. At the end of the game, there is a character | The swiping required to erase the journal gains layered meaning. Despite being a destructive act – and one's finger feeling like a clumsy blunt instrument in comparison to the delicate sketches – there is also a soothing quality to the gesture, which becomes more intense with repetition. It is akin |
|  |  | In moments of chaos and breakdown, the visuals become abstract, psychedelic, tessellating, jittery and confusing.<br>In the final scenes of restoration and relief, the colours of the sketched characters become deep and saturated, making them feel more 'real' than the simple outlines. | neon static. At this point, the electronic soundtrack becomes urgent, dissonant, and pulsating.<br>At the game's close, a woman sings lyrics that reflect the game's conclusion. | customisation activity and a chance for the player to insert their own name at the beginning of the journal. | to wiping the glass clean in order to see better, or stroking a friend to comfort them. |
| **What Remains of Edith Finch** | Following the death of her last living relative, a young girl returns to her ancestral home to learn the truth about a family curse. | The sinister, strangely-shaped ancestral home is a visual metaphor for the different generations of the Finch family. Each floor represents a new generation, and it teeters into a precarious tower, with Edith's bedroom located right at the top. The power of storytelling is prominent in the visuals – the house itself is filled with books that spill from shelves and tower in stacks on the floor. Different media trigger leaps into ancestral bodies, including a comic strip, a photo album, and even some legal divorce papers. | The pretty but ominous music box theme song reflects the entwined stories of childhood mortality. It is both whimsical and melancholic. | The protagonist temporarily inhabits the bodies of her ancestors to live out the moments before their deaths. These are often magical realist in nature – for example, the death of 7-year-old Molly is represented as her changing from one animal form into another until she becomes a carnivorous sea monster that waits under her own bed to devour her sleeping self. | Each time Edith inhabits a new ancestral body, the controls change slightly, meaning that both the player and Edith have to 'familiarise' themselves with a new schema. This mirrors the idea of 'getting to know' a deceased relative through embodied empathy. |
| **Röki** | A young girl processes the loss of her mother whilst rescuing her little brother from a folkloric monster. | The folkloric monsters Rorka and Roki represent 'shadow versions' of Tove's mother and brother. Since Tove's mother died birthing her brother, Rorka is depicted sacrificing herself to turn her monstrous son into a human being.<br>The giant sleeping Jotun who are meant to guard the forest represent Tove's grieving father who is passed | When Tove is forced to confront her repressed memory of her mother's death, she is haunted by the sound of a car's engine failing to start and the endless ringing of the emergency payphone. Tove was sent into the | Tove collects items to solve environmental puzzles. She keeps the items in her rucksack and can drag and drop them together to combine them. She also collects souvenirs that represent her repressed memories. | When Tove has to face the guilt she feels over her mother's death, the player has to hold the 'up' key for an extended period to have Tove run away from the player and towards the accusatory head of Rorka, the shadow-mother. |
|  |  | out in an alcohol-induced slumber at the time of her little brother's abduction. | forest to use the payphone to call for help but couldn't find in time, and so blames herself for her mother's death. | In the penultimate sequence of the game, Tove and her father must work cooperatively to solve environmental puzzles, and the player can switch between characters. |  |
| **Knights and Bikes** | A child grieving her mother's death seeks a mythical treasure to prevent her father's eviction by a predatory landlord, following clues left in her mother's journal. | The hand painted, mixed-media visual style evokes the creative output of a primary school collage project. It combines scribbly, messy doodles with the textures of crepe paper, cardboard, and playdough. The Arthurian legend of the cursed treasure often looks less 'real' than other visual elements – it is usually just drawn as an outline without depth or shading. | The soundtrack is a nostalgic homage to adolescent 80s punk. The sound effects often endorse the child protagonists' imaginative play – for example, their bikes make realising horse sounds when they are cycling out on adventures. | Co-operative mechanics are required for solving environmental puzzles and defeating enemies in combat. Nessa's ranged attacks (with a frisbee) complement Demelza's melee attacks (with her wellies). Healing uses a shared resource of plasters, and requires one player to hold a button and wait for the other player to approach them and press the corresponding button to give them a healing hi-five. | A sense of free-wheeling fun is created through the slightly chaotic combat and the informal 'races' that emerge when exploring the island. Interdependence is firmly established between the protagonists as players instinctively synchronise their input and keep one eye on each other's progress. |
| **Lost Words: Beyond the Page** | A child navigates the loss of her beloved grandmother. | Hand drawn illustrations and handwritten text in the journal – the sketches turn grey following the grandmother's death. When the player enters the imaginative world of the young writer's fiction (Estoria), the game becomes a fully coloured vibrant side-scrolling world. The fictional heroine must fight an unbeatable dragon that represents death. | Fully voice acted. Protagonist expresses grief through tone and pace of speech. There is an orchestral soundtrack that reinforces moments of emotional climax. | The player navigates a 2D platformer set within the protagonist's journal, using words as platforms and rearranging select words to advance. The heroine's journey through Estoria reflects the five stages of grief, with characters like a Djinn for denial and a giantess for anger representing each | The controls can feel fiddly. Having to execute the platforming with one hand on the keyboard and whilst moving the magical verb around with other hand on the mouse feels clumsy. |
|  |  | The heroine must collect dispersed fireflies to unlock their restorative power. |  | stage. She gains magical verbs (e.g., 'raise,' 'repair') to solve puzzles, symbolising her emotional growth and coping mechanisms. |  |
| **Under a Star called Sun** | The protagonist records daily life in a space station, including the use of a machine that allows them to relive their last memory of a loved one. | As the protagonist replays their last memory of the deceased over and over, the file becomes more corrupted and is distorted with glitches and digital artefacts. There is a repeated reference to the perseverance of Atlas, holding up the firmament knowing he has no real power to change things. | The 8-bit audio starts as a drudging monotonous tone, which is then layered with a more upbeat melody. | Beyond walking around the space station, the only mechanics are pressing the forward arrow to progress the message, reflecting the unchanging trajectory of the spaceship away from earth. | The monotony of daily life without the loved one is reflected in the simple, repetitive controls. The inexorable progression of time is felt in the temporal leaps forward triggered by a single button press. |

**Table 3.**
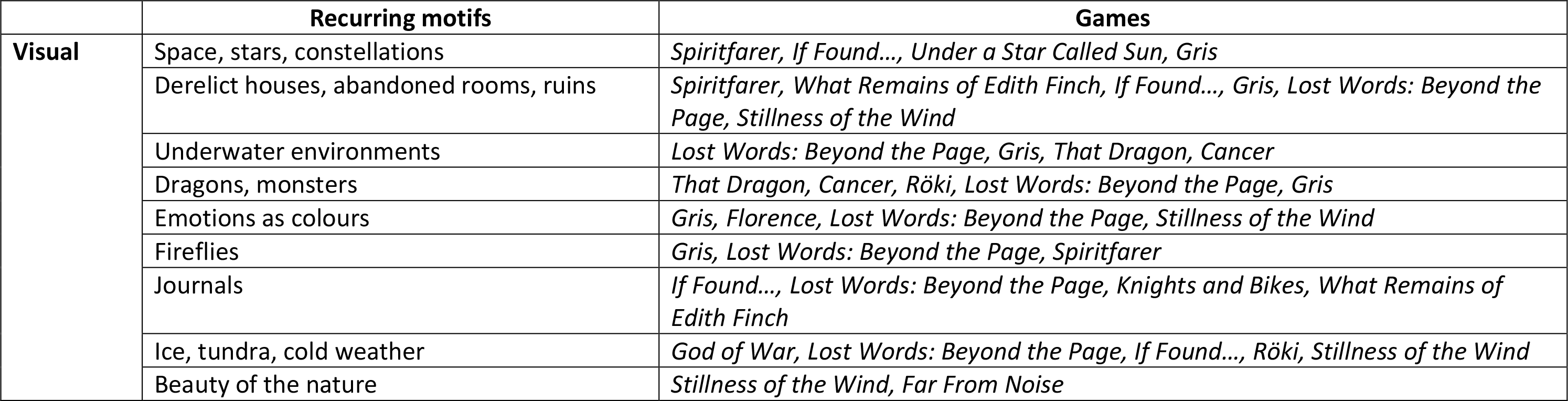
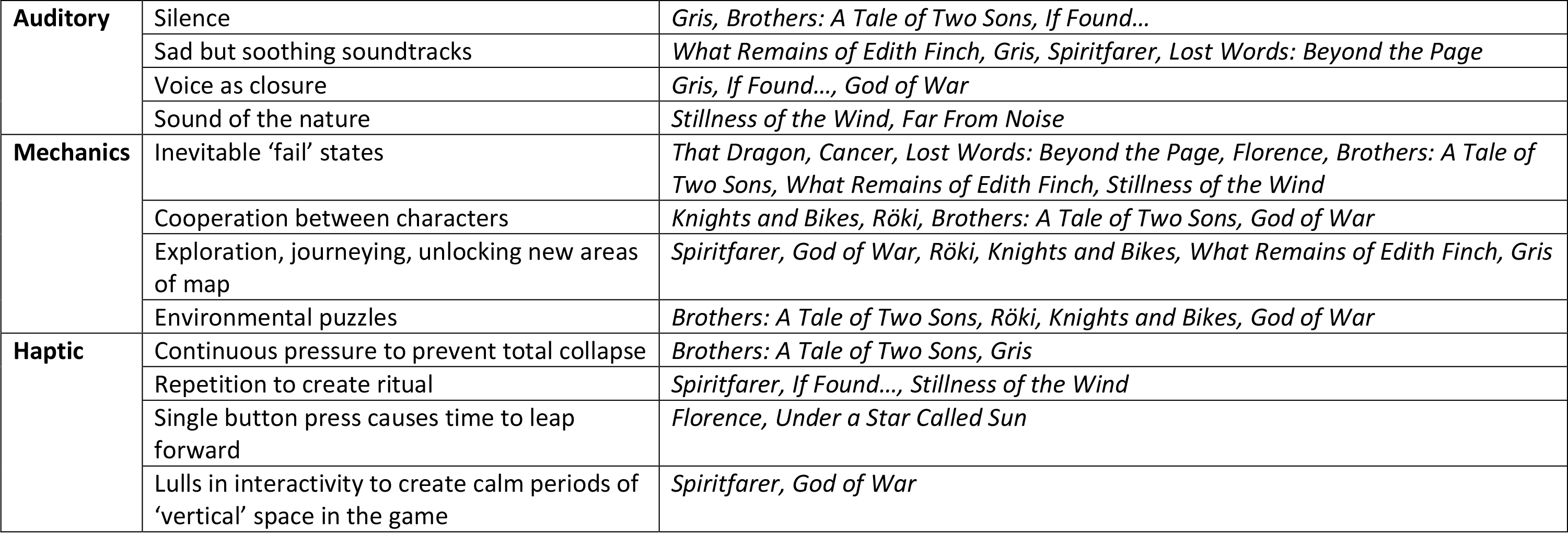
Recurring motifs within the selected games.

|  | Recurring motifs | Games |
| --- | --- | --- |
| **Visual** | Space, stars, constellations | *Spiritfarer, If Found..., Under a Star Called Sun, Gris* |
|  | Derelict houses, abandoned rooms, ruins | *Spiritfarer, What Remains of Edith Finch, If Found..., Gris, Lost Words: Beyond the Page, Stillness of the Wind* |
|  | Underwater environments | *Lost Words: Beyond the Page, Gris, That Dragon, Cancer* |
|  | Dragons, monsters | *That Dragon, Cancer, Röki, Lost Words: Beyond the Page, Gris* |
|  | Emotions as colours | *Gris, Florence, Lost Words: Beyond the Page, Stillness of the Wind* |
|  | Fireflies | *Gris, Lost Words: Beyond the Page, Spiritfarer* |
|  | Journals | *If Found..., Lost Words: Beyond the Page, Knights and Bikes, What Remains of Edith Finch* |
|  | Ice, tundra, cold weather | *God of War, Lost Words: Beyond the Page, If Found..., Röki, Stillness of the Wind* |
|  | Beauty of the nature | *Stillness of the Wind, Far From Noise* |
| **Auditory** | Silence | *Gris, Brothers: A Tale of Two Sons, If Found...* |
|  | Sad but soothing soundtracks | *What Remains of Edith Finch, Gris, Spiritfarer, Lost Words: Beyond the Page* |
|  | Voice as closure | *Gris, If Found..., God of War* |
|  | Sound of the nature | *Stillness of the Wind, Far From Noise* |
| **Mechanics** | Inevitable 'fail' states | *That Dragon, Cancer, Lost Words: Beyond the Page, Florence, Brothers: A Tale of Two Sons, What Remains of Edith Finch, Stillness of the Wind* |
|  | Cooperation between characters | *Knights and Bikes, Röki, Brothers: A Tale of Two Sons, God of War* |
|  | Exploration, journeying, unlocking new areas of map | *Spiritfarer, God of War, Röki, Knights and Bikes, What Remains of Edith Finch, Gris* |
|  | Environmental puzzles | *Brothers: A Tale of Two Sons, Röki, Knights and Bikes, God of War* |
| **Haptic** | Continuous pressure to prevent total collapse | *Brothers: A Tale of Two Sons, Gris* |
|  | Repetition to create ritual | *Spiritfarer, If Found..., Stillness of the Wind* |
|  | Single button press causes time to leap forward | *Florence, Under a Star Called Sun* |
|  | Lulls in interactivity to create calm periods of 'vertical' space in the game | *Spiritfarer, God of War* |

## Discussion

The video games in our corpus could be positioned along sliding scales from ‘literal’ to ‘figurative’ and from ‘realistic’ to ‘fantastical’. *God of War*, for example, is quite literal in how it presents the mourning of its central characters: skilled actors give expert performances of the intrapsychic and interpersonal effects of grief through naturalistic dialogue and believable body language. However, this game is ‘fantastical’ in the sense that the plot centres of battles between Norse gods, mythical creatures, dwarves, witches, and legions of undead. *Gris,* on the other hand, is highly figurative. The nature of the loss itself is left ambiguous – some players interpret it as the death of a mother while others see it as the loss of a previously held identity. Some games had ‘literal’ and ‘realistic’ elements, but deeply metaphorical subplots. *If Found…*recounts a believable experience of a three-dimensional character in a specific time period and setting – rural Ireland in the early 90s.

This narrative is entwined with a sci-fi subplot about an intergalactic explorer fighting to destabilise a blackhole. Similarly, *Lost Words: Beyond the Page* shifts between the realistic story of a young girl losing her grandmother and the highly metaphorical world of Estoria – a fictional place invented by the girl as the backdrop for her first novel. However, even games in our corpus that took a literal, realistic approach to exploring grief, relied on multimodal metaphors to convey and elicit feelings associated with loss.

We identified a recurrent set of metaphors that appeared across multiple video games. Several games used images of outer space to describe loss. In *If Found…* and *Under a Star Called Sun,* outer space expresses the vastness of loneliness and the emptiness of loss. Hurtling away from Earth, alone and untouchable in the vacuum of space, the voyager in *Under a Star Called Sun* connects the inexorable passage of time to the growing emotional distance from a lost loved one. Rather than putting grief into perspective, the increasing distance seems to exacerbate the sense of loss because the destination of the spaceship is unclear and, perhaps, unimportant to the voyager. In *If Found…,* the sci-fi subplot follows the efforts of an intergalactic explorer to prevent a blackhole from engulfing the world by creating an ‘anomaly’. The anomaly occurs when Kasio’s mother is given a drawing an astronaut that Kasio made as a child, connecting the subplot with the main plot and making explicit the fact that the ‘blackhole’ is a metaphor for Kasio’s potential death through community- and self-neglect. *Spiritfarer* and *Gris* use visual metaphors of constellations to illustrate restorative forms of grieving. Metaphors of seeing light during moments of darkness undergird star-gazing practices of finding narrative meaning by joining previously unconnected dots. Completing these patterns allows the avatar in *Gris* to walk new pathways through the sky, while the constellations that appear in *Spiritfarer* act as reminders that although the deceased are beyond our reach, they persist in new cosmic forms. The collection mechanic associated with fireflies in multiple games also draws on metaphors of focussing on glimmers of light during dark times, even if those lights are faint, transient, and fugitive.

The images of ruins, abandoned houses, and bedrooms as proxies for deceased characters appeared in over a third of the games in our corpus. The gothic ‘haunted house’ trope is a very common metaphor for unprocessed grief, but is perhaps especially prevalent in video games because this medium often relies on environmental storytelling to impart narrative (Jenkins 2004). In *What Remains of Edith Finch,* the house itself is a metaphor for family. It expresses both the welcoming hospitality of family and the dangerous, jealous claustrophobia of overbearing relatives. *What Remains of Edith Finch* models the concept of ‘getting to know’ lost loved ones by engaging with artefacts associated with them and with stories told about them. In *If Found…* and *Gris* the crumbling structures represent the decline of the protagonists’ wellbeing and sense of one’s identity becoming fragmented and fractured as grief erodes one’s firm foundation. In *Spiritfarer,* the abandoned room takes on a slightly more positive meaning. The empty bedrooms of former passengers are a constant reminder of those who have passed on and their material persistence is juxtaposed to the immateriality of their former occupants. However, there is something comforting about being able to revisit the spaces that capture some of the essence and personality of the dead that is not unlike the comfort of visiting a gravestone.

The ‘depression as drowning’ analogy is also familiar from other media. However, in its deployment in the games in our corpus, the overwhelm conveyed by waves of sadness were counterbalanced by an opening up of exploration opportunities. The undersea caverns in *Gris,* for example, offer players new directions for discovery, while the ocean in *That Dragon, Cancer* represents a change from the safe but strictly delimited island environment. In this sense, the idea of ‘wallowing in one’s sadness’ seems less pejorative since the embodied metaphor acknowledges the allure and perhaps even the importance of ‘plumbing the depths’ of one’s depressive feelings. Similarly, the cathartic release associated with rage – for instance, the ‘juicy’ combat of *God of War* or the sense of destructive release associated with smashing vases in *Gris* – means that this emotion is not cast as something ‘negative’ that must be avoided or moved past. The free movement associated with the fluidity of water is contrasted to the numbing paralysis of ice. In *Röki,* the final challenges take place in a palace of ice, and in *God of War* Kratos must learn to master the cold of emotional repression and the burning heat of rage before he can achieve self-acceptance.

Representing illness and death as monsters is not unique to video games, but when coupled with mechanics that force fail states these metaphors serve to undermine the ‘battle / soldier’ rhetoric attached to terminal diagnoses. Due to the strong conventional association between failure and death in video games, games in our corpus, such as *That Dragon, Cancer, Lost Words: Beyond the Page, Florence, Brothers: A Tale of Two Sons, What Remains of Edith Finch, Stillness of the Wind*, were able to subvert this association by suggesting that overcoming death is not possible through skill, will, or effort.

Four games in our corpus, i.e., *If Found…, Lost Words: Beyond the Page, Knights and Bikes, What Remains of Edith Finch*, were framed by or set within the focalising the characters’ journals. The power of autobiographical storytelling as a grieving practice.

Many games achieved feelings of intimacy using art styles and fonts that were suggestive of being hand drawn and handwritten. Since grieving can be a vulnerable and personal experience, these design decisions add a ‘human thumbprints’ to the digital text, affirming the presence and investment of human creators.

Beyond visual and auditory representation of grief, the use of game mechanics and haptic offers a novel approach to influencing player emotions. For instance, *Florence* utilises jigsaw puzzle difficulty to mirror the challenges of communication within a relationship’s various phases. Initially, puzzles are more complex, representing the couple’s cautious and deliberate communication. As their relationship deepens, the puzzles simplify, reflecting improved understanding and ease of communication. Moreover, the game introduces a mechanic where players must restrain their interaction to symbolise the need to “let go” when the relationship ends. In *Stillness in the Wind*, the game’s pace intentionally slows down, symbolising the protagonist’s aging process. This mechanic not only adds depth to gameplay but also profoundly conveys time’s passage, inevitable change, and the exploration of life, death, loneliness, and despair.

## Conclusion and Limitations

This article traverses the intersection of games studies, medical ethics, and clinical psychology to dissect how contemporary video games metaphorically represent grief. It illuminates the multifaceted nature of grief—a universally yet complexly navigated experience—and highlights how video games, through their unique blend of audio, visual, verbal, haptic elements, and game mechanics, offer new metaphors for articulating and exploring the experience of loss. This exploration covers 14 COTS games, identifying recurring motifs, their illumination and obscuration of grief, and their therapeutic potentials. Ultimately, the paper proposes recommendations for game developers aiming to tackle emotionally charged topics in serious games and for therapists interested in incorporating games into grief counselling. This multidisciplinary approach not only broadens the understanding of video games’ potential in therapeutic contexts but also challenges traditional narratives surrounding grief, suggesting that the medium’s inherent interactivity and multimodality could redefine the paradigms of grief therapy.

The paper suggests that video games’ capacity to evoke a spectrum of emotions through gameplay mechanics, including discomfort and ambiguity, positions them uniquely for addressing themes of grief. It demonstrates the role of games in facilitating a nuanced exploration of loss, beyond merely softening the edges of difficult emotions. We have shown that metaphoric representation creates interpretive gaps that encourage the player to co-construct meaning. Recommendations for game developers focus on leveraging the interactive and participatory nature of games to enable players to explore grief in a manner that is personal and meaningful. These insights advocate for a nuanced approach to designing serious games, where the complexity of human experiences like grief can be approached with empathy, creativity, and a readiness to embrace interpretive gaps for players to express their own meaning through choice and interaction. This perspective champions video games as a significant avenue for therapeutic exploration, heralding a new chapter in how we perceive and utilise games in the mental health and healing.

However, it is crucial to acknowledge the limitations related to cultural perspectives on grief. Variations in how different cultures process grief—from specific mourning periods to annual remembrance days like the Qingming festival (a.k.a. tomb sweeping day) in Chinese culture—highlight the contextual and cultural embeddedness of grief metaphors. Our inclusion criterion of games available in English limits the cultural perspective to primarily Western interpretations of grief. Metaphors, deeply ingrained in cultural constructs, vary across cultures, social groups, countries, and identity groups. As Littlewood (1990) argues, context dependent meanings are more important than specific universal processes of healing. Thus, video games may introduce metaphors that do not universally translate across cultural or social divides, suggesting a need for broader inclusivity and consideration in future research.

## Data Availability

All data produced in the present work are contained in the manuscript

## Acknowledgement

This work was funded by the MRC/AHRC/ESRC Adolescence, Mental Health and the Developing Mind Programme (Project name: ATTUNE—Understanding mechanisms and mental health impacts of Adverse Childhood Experiences to co-design preventive arts and digital interventions. Grant number: MR/W002183/1) and supported by the NIHR Applied Research Collaboration Oxford and Thames Valley at Oxford Health NHS Foundation Trust. We would like to thank the following individuals for their valuable contributions in acquiring research funding, project management, discussions, editing and proofreading: Anton Belinskiy, Siobhan Hugh-Jones, Tanya Krzywinska, Paul McCrone, Sania Shakoor, Mina Fazel, Isabelle Butcher, Simran Sansoy, Paul Cooke, Nick Smith, David Prior and Douglas Brown.

## References

Barbara, J. (2020). Twine and DooM as Authoring Tools in Teaching IDN Design of LudoNarrative Dissonance. In A.-G. Bosser, D. E. Millard, & C. Hargood (Eds.), Interactive Storytelling (pp. 120–124). Springer International Publishing. 10.1007/978-3-030-62516-0_11

Bogost, I. (2007). Persuasive Games: The Expressive Power of Videogames. MIT Press.

Gee, J. P., & Hayes, E. R. (2011). Language and Learning in the Digital Age. Routledge.

Juul, J. (2009). A Casual Revolution: Reinventing Video Games and Their Players. MIT Press.

Carr, D. (2019). Methodology, Representation, and Games. Games and Culture, 14(7–8), 707–723. 10.1177/1555412017728641

Caserman, P., Hoffmann, K., Müller, P., Schaub, M., Straßburg, K., Wiemeyer, J., Bruder, R., & Göbel, S. (2020). Quality Criteria for Serious Games: Serious Part, Game Part, and Balance. JMIR Serious Games, 8(3), e19037. 10.2196/19037

Deterding, S. (2020). Cookie Clicker: Gamification (Vol. 1, pp. 200–207). New York, USA: New York University Press. 10.18574/9781479830404-026

Eum, H., Chen, C., Rosson, M. B., & Carroll, J. M. (2021). Digital mourning: The evolution of mourning practices in digital spaces. In Proceedings of the 2021 CHI Conference on Human Factors in Computing Systems.

Eum, K., Erb, V., Lin, S., Wang, S., & Doh, Y. Y. (2021). How the Death-themed Game Spiritfarer Can Help Players Cope with the Loss of a Loved One. Extended Abstracts of the 2021 CHI Conference on Human Factors in Computing Systems, 1–6. 10.1145/3411763.3451608

Farber, M., & Schrier, K. (2021). Beyond Winning: A Situational Analysis of Two Digital Autobiography GAmes. Game Studies, 21(4). http://gamestudies.org/2104/articles/farber_schrier

Fazel, M., Stratford, H. J., Rowsell, E., Chan, C., Griffiths, H., & Robjant, K. (2020). Five Applications of Narrative Exposure Therapy for Children and Adolescents Presenting With Post-Traumatic Stress Disorders. Frontiers in Psychiatry, 11. https://www.frontiersin.org/articles/10.3389/fpsyt.2020.00019

Fitzgerald, M., & Ratcliffe, G. (2019). Serious Games, Gamification, and Serious Mental Illness: A Scoping Review. Psychiatric Services, appips201800567.

Flanagan, M. (2009). Critical play: Radical game design. MIT Press.

Freeman, D. (2004) Creating emotion in games: The craft and art of emotioneering™, Comput. Entertain. (CIE), 2(3), p.15

Harrer, S. (2013). From Losing to Loss: Exploring the Expressive Capacities of Videogames Beyond Death as Failure. Culture Unbound: Journal of Current Cultural Research, 5, 607– 620team. 10.3384/cu.2000.1525.135607

Harrer, S. (2018). Games and Bereavement. transcript Verlag.

Harrer, S., & Schoenau-Fog, H. (2015). Inviting Grief into Games: The Game Design Process as Personal Dialogue. DiGRA Conference.

Hocking, C. (2007, October). Ludonarrative Dissonance in Bioshock. Click Nothing. https://clicknothing.typepad.com/click_nothing/2007/10/ludonarrative-d.html

Jowett, B. (n.d.). Republic, Book X. Retrieved from http://classics.mit.edu/Plato/republic.11.x.html

Karlsen, F. (2022). Balancing Ethics, Art and Economics: A Qualitative Analysis of Game Designer Perspectives on Monetisation. Games and Culture, 17(4), 639–656. 10.1177/15554120211049579

Karlsen, F. (2022). Mobile games and the app store: The impact of market saturation on game design. Journal of Gaming & Virtual Worlds, 14(1), 45–59.

Kłosiński, M. (2022). How to Interpret Digital Games? A Hermeneutic Guide in Ten Points, With References and Bibliography. Game Studies, 22(2). http://gamestudies.org/2202/articles/gap_klosinski+

Kłosiński, R. (2022). “Digital games as autonomous aesthetics: toward a post-positivist game studies.” Games and Culture. doi: 10.1177/15554120211008633

Kübler-Ross, E. (2014a). On death & dying: What the dying have to teach doctors, nurses, clergy & their own families / Elisabeth Kübler-Ross (Scribner trade paperback edition.). New York: Scribner, a division of Simon & Schuster, Inc., 2014.

Kübler-Ross, E. (2014b). On grief & grieving: Finding the meaning of grief through the five stages of loss / Elisabeth Kübler-Ross & David Kessler. (Anniversary edition.). London : Simon & Schuster, 2014.

Littlewood, R. (1990) How Universal is Something we can call ‘Therapy’? Some Implications of Non-Western Healing Systems for Intercultural Work, Holistic Medicine, 5(2), 49–65, DOI: 10.3109/13561829009043447

Munsterburg, M. (n.d.). “Writing about Art: Ekphrasis.” Retrieved from http://www.writingaboutart.org/pages/ekphrasis.html

Nakamura, L. (2002). Cybertypes: Race, ethnicity, and identity on the Internet / Lisa

Nakamura. Place of publication not identified : Routledge.

Nicklin, H. (2022). Writing for games: Theory and practice / Hannah Nicklin. (1st.).

Pozo, T. (2018). Queer Games After Empathy: Feminism and Haptic Game Design Aesthetics from Consent to Cuteness to the Radically Soft. Game Studies, 18(3).

Răzman, D. C. (2021). Press ‘F’ to pay respects: Grief and memorialization in video games. http://urn.kb.se/resolve?urn=urn:nbn:se:his:diva-20098

Răzman, M. A. (2021). Dealing with Death in Mobile Games. Romanian Journal of Journalism and Communication, 18(1), 1–16.

Reay, E. (2018). Appraising the Poetic Power of Children’s Video Games. International Research in Children’s Literature, 11(1), 17–32. 10.3366/ircl.2018.0251

Robjant, K., & Fazel, M. (2010). The emerging evidence for Narrative Exposure Therapy: A review. Clinical Psychology Review, 30(8), 1030–1039. 10.1016/j.cpr.2010.07.004

Ruberg, B., & Scully-Blaker, R. (2021). Making players care: The ambivalent cultural politics of care and video games. International Journal of Cultural Studies, 24(4), 655–672. 10.1177/1367877920950323

Rusch, D. (2009). Mechanisms of the Soul–Tackling the Human Condition in Videogames.

Sexton, S. (2019). If all the world and love were young / Stephen Sexton. In If all the world and love were young.

Steirer, G., & Barnes, N. G. (2019). Four Components of Mobile Gaming: Gameplay Accessibility, Software Accessibility, Everyday Ubiquity, and Variable Monetization. Games and Culture, 14(3), 217–237.

Stieler-Hunt, C., Jones, C., Rolfe, B., & Pozzebon, K. (2014). Examining key design decisions involved in developing a serious game for child sexual abuse prevention. Frontiers in Psychology, 5. https://www.frontiersin.org/article/10.3389/fpsyg.2014.00073

Stone, J. (2021). Separation Anxiety: Plotting and Visualising the Tensions Between Poetry and Videogames. Game Studies, 21(2).

Sundaram, D., & Gottlieb, O. (2022). “It’s so normal, and … meaningful.” Playing with Narrative, Artifacts, and Cultural Difference in Florence. Gamevironments, 16, 68–99. 10.48783/gameviron.v16i16.185

Tekinbaş, K. S., & Zimmerman, E. (2006). The game design reader: A Rules of play anthology / [edited by] Katie Salen and Eric Zimmerman. Cambridge, Mass.

Trammell, A. (2020). Torture, Play, and the Black Experience. GAME: The Italian Journal of Game Studies, 8(9). https://www.gamejournal.it/torture-play/

Trammell, A. (2022). Decolonizing play. Critical Studies in Media Communication, 39(3), 239–246.

